# Involvement of mutant and wild-type *CYSLTR2* in the development and progression of uveal nevi and melanoma

**DOI:** 10.1101/2020.09.16.20191791

**Authors:** Rogier J. Nell, Nino V. Menger, Mieke Versluis, Gregorius P.M. Luyten, Robert M. Verdijk, Michele C. Madigan, Martine J. Jager, Pieter A. van der Velden

## Abstract

**Background:** Activating Gα_q_ signalling mutations are considered an early event in the development of uveal melanoma. Whereas most tumours harbour a mutation in *GNAQ* or *GNA11, CYSLTR2* (encoding G-protein coupled receptor CysLT_2_R) forms a rare alternative. The role of wild-type CysLT_2_R in uveal melanoma remains unknown.

**Methods:** We performed a digital PCR-based molecular analysis of benign choroidal nevi and primary uveal melanomas. Publicly available bulk and single cell sequencing data were mined to further study wild-type and mutant *CYSLTR2*.

**Results:** 1/16 nevi and 2/120 melanomas carried the *CYSLTR2* mutation. The mutation was subclonal in the nevus, while being clonal in both melanomas. In the melanomas, secondary, subclonal *CYSLTR2* alterations shifted the allelic balance towards the mutant. The resulting genetic heterogeneity was confirmed in distinct areas of both tumours. At the RNA level, further silencing of wild-type and preferential expression of mutant *CYSLTR2* was identified, which was also observed in 2/3 *CYSLTR2* mutant melanomas from the TCGA cohort. In *CYSLTR2* wild-type melanomas, high expression of *CYSLTR2* originated from melanoma cells and correlated to tumour inflammation.

**Conclusions:** Our findings suggest that *CYSLTR2* is involved in both early and late development of uveal melanoma. Whereas the *CYSLTR2* p.L129Q mutation is likely to be the initiating oncogenic event, various mechanisms further increase the mutant allele abundance during tumour progression. This makes mutant CysLT2R an attractive therapeutic target in uveal melanoma. In *GNAQ, GNA11* and *PLCB4* mutant melanomas, expression of wild-type *CYSLTR2* possibly facilitates an interaction with immune cells in the microenvironment and may also have therapeutic potential.

## Background

Uveal melanoma is the most common primary tumour in the adult eye, with an annual incidence of 2-8 per million (1). Up to ~50% of the patients develops metastatic disease, predominantly to the liver. After hepatic spreading, uveal melanoma is usually fatal within a year (2).

The cancer originates from melanocytes in the uvea, the pigmented vascular layer of the eye. It is hypothesised that (epi)genetic alterations drive the transformation of these melanocytes into benign nevi and subsequently into malignant melanomas. However, it is rare to find histologically distinct pre-malignant lesions within or adjacent to a primary tumour (3).

Activating Gα_q_ signalling mutations at mutually-exclusive hotspots in *GNAQ* and *GNA11* (p.Q209 and p.R183) are found in a large majority of uveal melanomas (4, 5). Their clonal abundance in primary tumours suggests that these mutations are acquired early in tumour development (6). The presence of these mutations in the majority of choroidal nevi supports this hypothesis (7).

The essential role of Gα_q_ signalling in uveal melanoma tumorigenesis has been further supported by the discovery of rare but recurrent mutations in *PLCB4* (p.D630) and *CYSLTR2* (p.L129Q) (8, 9). These genes, encoding phospholipase C (34 and a Gα_q_-coupled receptor respectively, are typically mutated in *GNAQ* and *GNA11* wild-type tumours, providing an alternative way of activating the Ga_c_ signalling pathway (10, 11).

*CYSLTR2* encodes for the G protein-coupled receptor cysteinyl-leukotriene receptor 2 (CysLT_2_R) which is, under wild-type conditions, involved in leukotriene-mediated signalling (12). Although CysLT_2_R has several well-studied functions (e.g. in alveolar inflammation) and can be expressed by a variety of cell types (e.g. eosinophils, basophils, neutrophils and macrophages), its natural role and origin in uveal melanomas remains unclear (13). Mutant *CYSLTR2* however leads to constitutive activation of endogenous Gα_q_ signalling, thereby stimulating the same pathways as oncogenic mutations in *GNAQ* and *GNA11* (10, 11).

In this study, we examined the presence of *CYSLTR2* mutations in non-malignant choroidal nevi and primary uveal melanomas. By targeted analysis using digital PCR, an accurate quantification of both *CYSLTR2* mutant and wild-type alleles could be achieved even in DNA samples of lower quality and quantity. In the *CYSLTR2* mutant melanomas, we further assessed the genetic heterogeneity within tumours and investigated the allelic balance of *CYSLTR2* at the RNA level. Expansion into publicly available bulk and single-cell uveal melanoma sequencing data allowed to study the role of wild-type *CYSLTR2* in melanomas with a mutation in *GNAQ, GNA11* or *PLCB4* (13, 14).

## Methods

### Sample collection and DNA/RNA extraction

As described earlier, 16 choroidal nevi were dissected from 13 formalin-fixed post-mortem human donor eyes, with consent from Lions NSW Eye Bank and approval from the University of Sydney and University of New South Wales Human Research Ethics Committee (7). The tissues were paraffin-embedded and 7 μm sections were cut and collected on SuperFrost Plus™ slides (Menzel-GlaLser, Braunschweig, Germany), dried and stored prior to use for routine haematoxylin and eosin staining. DNA was extracted using the ReliaPrep FFPE (Formalin-Fixed Paraffin-Embedded) gDNA Miniprep System (Promega Corp, Madison, USA).

120 fresh-frozen tumour samples, obtained from primary uveal melanomas treated by enucleation between 1999 and 2011, were available from the Department of Ophthalmology, Leiden University Medical Center (LUMC). All patient samples were collected with informed consent and this study was approved by the LUMC Biobank Committee and Medisch Ethische Toetsingscommissie under no. B14.003/DH/sh and B20.026. DNA and RNA were isolated from 25×25μm sections using the QIAamp DNA Mini Kit and RNeasy Mini Kit respectively (Qiagen, Hilden, Germany). cDNA was prepared from RNA using the iScript™ cDNA Sythesis Kit (Bio-Rad Laboratories, Hercules, USA). Total nucleic acid from microdissected tissue was isolated using the Siemens Tissue Preparation System (Siemens Healthcare, Erlangen, Germany) (15).

For all procedures, the manufacturer’s instructions were followed.

### Digital PCR

Digital PCR experiments were carried out using the QX200™ Droplet Digital™ PCR System (Bio-Rad Laboratories) following the protocols described earlier (6, 16, 17). A methodical overview of all experimental setups is presented in **Supplementary Data 1**. Context sequences, PCR annealing temperatures and supplier information for all assays used are provided in **Supplementary Table 1**.

Raw digital PCR results were acquired using *QuantaSoft* (version 1.7.4, Bio-Rad Laboratories) and imported in the online digital PCR management and analysis application *Roodcom WebAnalysis* (version 1.9.4, available via https://webanalysis.roodcom.nl).

### Sequencing

To validate the presence of the *CYSLTR2* mutation, 20 ng DNA was analysed in 7 uL experiment, using 3.5 uL iQ SYBR Green Supermix (Bio-Rad Laboratories) and primers in a final concentration of 500 nM. The PCR was performed in a CFX384 Real-Time PCR system (Bio-Rad Laboratories), using the same protocol as for digital PCR. Raw PCR products were purified and analysed for ‘Quick Shot Short’ Sanger Sequencing at BaseClear, Leiden, The Netherlands. AB1-files were analysed using *Roodcom SangerSeq Analysis* (version 1.0, https://sangerseq.roodcom.nl).

### TCGA data analysis

Segmented and arm-level SNP array copy number data files were downloaded from the Broad GDAC Firehose (https://gdac.broadinstitute.org). Aligned whole exome and RNA-sequencing files were obtained from the NCI Genomic Data Commons data portal (GDC; https://portal.gdc.cancer.gov). Heterozygous SNP’s were identified using freebayes (v1.3.1) (18). Allele-specific RNA expression was determined based on the balance between the *CYSLTR2* mutation versus wild-type and the balance between variants from 6 common SNP’s in the transcribed *CYSLTR2* gene. For both, the number of reads mapping to the specific variants were determined using samtools (v1.7) and freebayes (v1.3.1) (18, 19).

### Single cell data analysis

Processed sequencing data from 8 primary melanomas studied by Durante et al. (GSE139829) was downloaded from the Gene Expression Omnibus repository (www.ncbi.nlm.nih.gov/geo/cuery/acc.cgi?acc=GSE139829) (14). The data set was analysed in R (v3.6.0) using the Seurat library (v3.1.4), following the methodology from the original study (20). Uveal melanoma cells were identified based on gene expression of *MLANA* (Melan-A). Clustering was conducted using 10 principle components and a resolution parameter set to 0.5, and was visualized in a two-dimensional tSNE plot.

### Code availability

All custom scripts used in this study are available via https://git.lumc.nl/rjnell/cysltr2.

## Results

### *CYSLTR2* p.L129Q is subclonally present in a *GNAQ/GNA11* wild-type nevus

Previously, 16 choroidal nevi from 13 post-mortem collected eyes were analysed for *GNAQ* and *GNA11* hotspot mutations. In 15 cases such mutations were found, but always in a subfraction of the nevus cells (7). In the same cohort we applied a custom digital PCR assay, which targets the *CYSLTR2* p.L129Q wild-type and mutant allele. In nevus 12B, the only one being *GNAQ* and *GNA11* wild-type, we now identified this *CYSLTR2* mutation (**Figure 1**).

**Figure 1.**
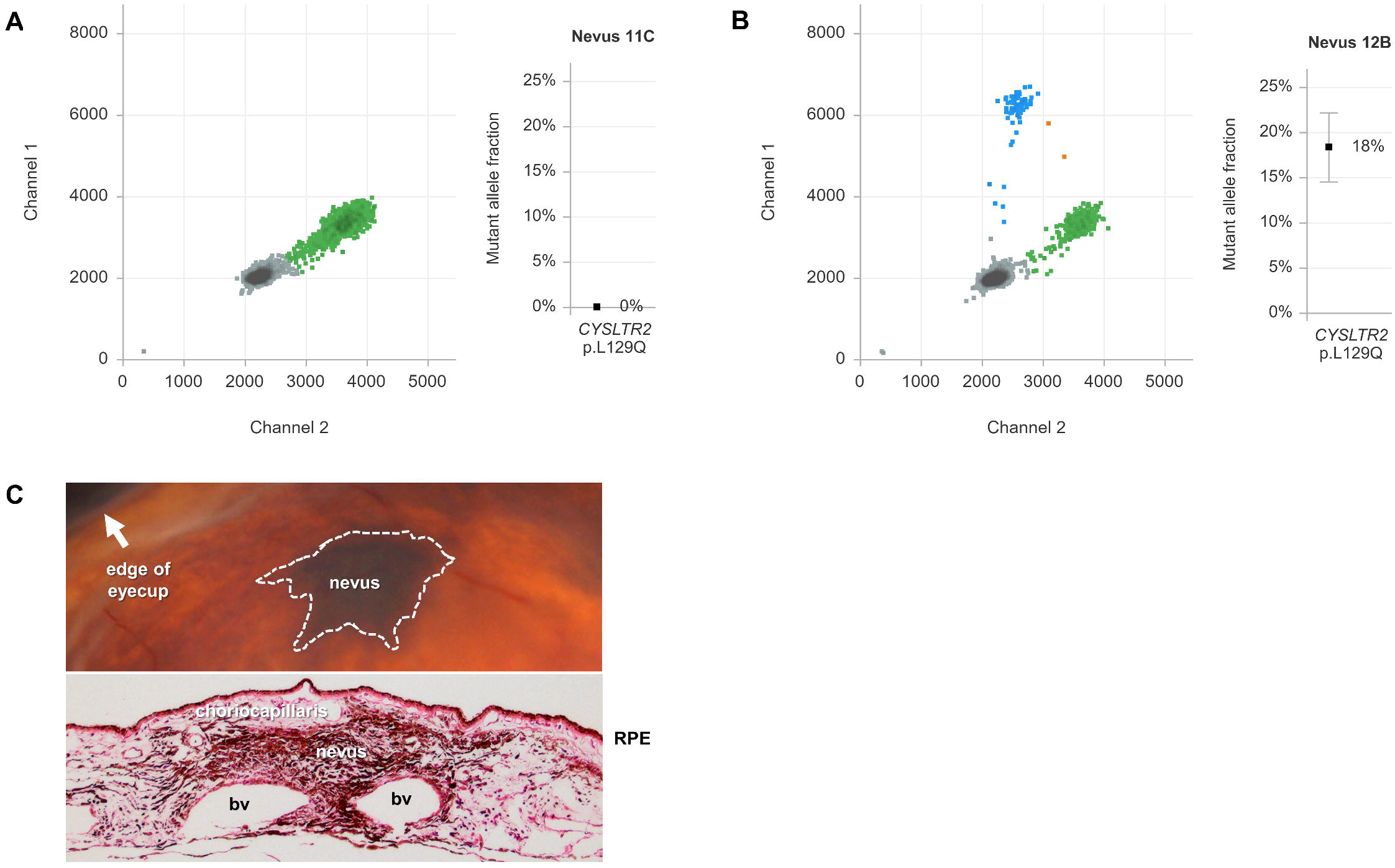
Analysis of benign choroidal nevi. (**A**) Duplex analysis of *CYSLTR2* p.L129Q mutation in choroidal nevus 11C using digital PCR. The 2D plot at the left illustrates that no droplets are positive for *CYSLTR2* mutant alleles on channel 1 (FAM labelled, in blue) while *CYSLTR2* wild-type alleles are abundantly detected on channel 2 (HEX labelled, in green). Empty droplets (in grey) do not contain the *CYSLTR2* target amplicon. At the right side the calculated mutant allele fraction with 95% confidence interval confirms the absence of mutant alleles. (**B**) Similar analysis for *CYSLTR2* p.L129Q mutation in nevus 12B, with detectable mutant alleles on channel 1 (FAM labelled, in blue). Double-positive droplets (in orange) contain both a mutant and wild-type allele. The mutant allele fraction is calculated to be 18.4%. (**C**) Upper panel: in-eye image showing nevus 12B as a dark choroidal lesion, with arrow indicating the eye cup. Lower panel: haematoxylin and eosin staining showing the nevus as highly pigmented lesion in the choroid. RPE = retinal pigment epithelium, bv = blood vessel.

The mutant allele fraction of *CYSLTR2* p.L129Q in this nevus was 18%, representing a mutant cell fraction of 36%. As an earlier Melan-A staining indicated a >95% nevus cell purity of the macrodissected tissue, it turns out that only a subclonal fraction of the nevus cells was mutated (**Figure 1C**) (7). Nevertheless, we were able to validate the mutation by Sanger sequencing (**Figure S1**).

### *CYSLTR2* p.L129Q is found in 2/120 primary uveal melanomas

A cohort of 120 archival primary uveal melanomas, treated by enucleation at the LUMC, was screened for the *CYSLTR2* p.L129Q mutation. This mutation was found in 2 tumours (~2%), which were both *GNAQ* and *GNA11* wild-type (**Supplementary Table 2**). Remarkably, both cases showed a mutant allele fraction significantly higher than 50%, being at odds with heterozygosity for the mutant allele (**Figure 2A**). To resolve the exact genetic composition and evolutionary history of these two melanomas, multiple spatially separated samples of both tumours were investigated using duplex and multiplex digital PCR (**Supplementary Data 1**).

**Figure 2.**
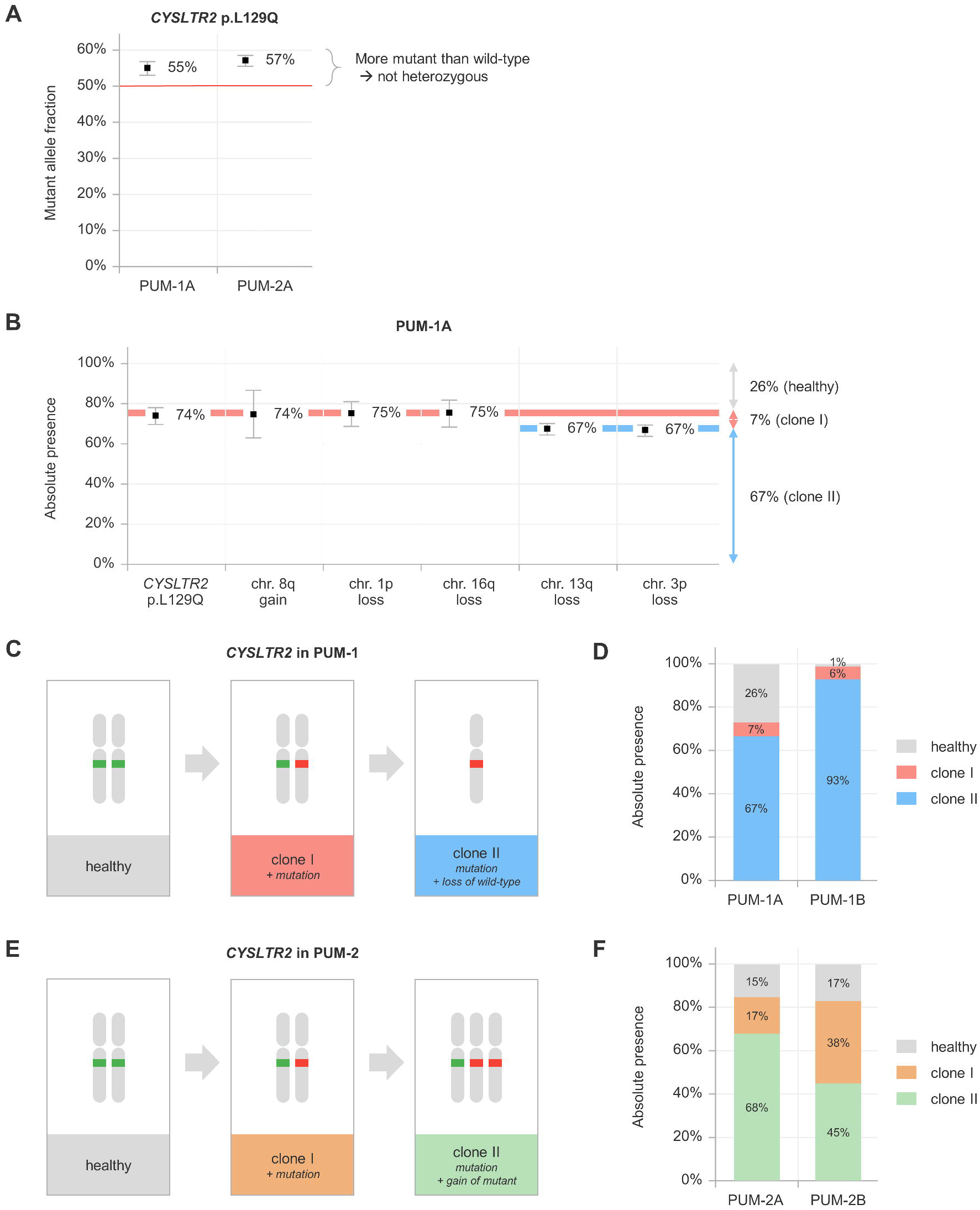
Characterisation and molecular evolution of *CYSLTR2* mutant uveal melanomas. (**A**) Mutant allele fractions for *CYSLTR2* p.L129Q in PUM-1A and PUM-2A are both significantly higher than 50% and therefore in conflict with heterozygosity of this mutation. (**B**) Absolute quantification of genetic alterations in PUM-1A. *CYSLTR2* mutation, gain of chromosome 8q and losses of chromosome 1p and 16q present being clonal in a fraction of ~74% of total cells in the tumour mass. The losses of chromosome 13q and chromosome 3p are subclonal, as both are detected in 67% of the cells. (**C**) *CYSLTR2* genetic evolution in PUM-1A/B, as inferred from absolute quantification of genetic aberrations. Starting at the germline situation (healthy cells), firstly the mutation has been acquired (clone I), after which the wild-type allele has been eliminated (clone II). (**D**) Clonal composition of PUM-1A/B shows the presence of both clones in both samples, but a lower fraction healthy cells is observed in the microdissected sample (PUM-1B: 1%) versus the macrodissected sample (PUM-1A: 26%). (**E**) *CYSLTR2* genetic evolution in PUM-2A/B, as inferred from absolute quantification of genetic aberrations. Starting from the germline situation (healthy cells), firstly the mutation has been acquired (clone I), after which the mutant allele has been gained (clone II). (**F**) Clonal composition of PUM-2A/B shows the presence of both clones in both samples at varying levels.

### Genetic composition of *CYSLTR2* mutant melanoma 1 (PUM-1)

PUM-1 was obtained from a woman (age in range 85-90 years) with a large uveal melanoma localised to the ciliary body of the right eye. Clinical characteristics of this case are presented in **Supplementary Table 2**. From this patient, one macrodissected (PUM-1A) and one microdissected (PUM-1B) area of the primary tumour were analysed.

Digital PCR for the *CYSLTR2* p.L129Q mutation in PUM-1A revealed a mutant allele fraction of 55% (**Figure 2A**). Using our multiplex digital PCR setup (**Supplementary Data 1**), we identified loss of the wild-type *CYSLTR2* allele (chromosome 13q) in 67% of the cells within the tumour mass (**Figure 2B**). This was confirmed by the allelic imbalance of heterozygous common single nucleotide polymorphism (SNP) rs2057413 in *SETDB2*, located ~776kb telomeric from *CYSLTR2* (**Supplementary Data 1**). Taking the chromosome 13q copy number alteration into account, the mutant cell fraction was calculated to be 74%. Additional digital PCR experiments showed that losses of 1p and 16q, and gain of 8q were also present in ~74% of the cells. However, loss of chromosome 3p was detected in 67% of the cells.

The comparable abundances (~74% of total tumour) of the *CYSLTR2* mutation, gain of 8q and losses of 1p and 16q suggest that these alterations are clonally present in this sample and occurred during an early developmental phase. The losses of both wild-type *CYSLTR2* and chromosome 3p were present in a significantly smaller, subclonal proportion of the cancer cells (67% of total tumour), which indicates that these alterations occurred later during tumour development and that these cells coexisted with the predecessor clone. Taken together, the two clones represented 7% and 67% of the complete tumour mass, while another 26% of non-malignant, healthy cells complemented the tumour composition (**Figure 2B-D**).

Analysis of a melanoma cell-rich area of the primary tumour, obtained by microdissection (PUM-1B), was specified to the single-amplicon assays *(CYSLTR2* mutation and SNP assays on chromosome 3p and 13q) to prevent biases due to degradation of FFPE-based input material. Again, two different clones were identified: in 99% of the cells the *CYSLTR2* mutation was present, with chromosome 3p and 13q lost in 93% of the cells, giving clonal sizes of 6% and 93%, and a healthy cell fraction of 1% (**Figure 2D**).

The genetic alterations of PUM-1 are typically associated with a poor prognosis, which is highlighted by the observed negative *BAP1* nuclear staining (**Supplementary Table 2**) (16). This patient died of unknown cause 20 months after enucleation.

### Genetic composition of *CYSLTR2* mutant melanoma 2 (PUM-2)

PUM-2 presented as an iridociliary melanoma in an male child (age in range 5-10 years), which led to enucleation of the right eye. An overview of the clinical characteristics is presented in **Supplementary Table 2**. From this patient, one macrodissected (PUM-2A) and one microdissected (PUM-2B) area of the tumour were analysed.

In PUM-2A a *CYSLTR2* p.L129Q mutant allele fraction of 57% was observed (**Figure 2A**), which is also at odds with heterozygosity. In this tumour, a gain of chromosome 13q caused the relative excess of the mutant alleles (**Figure 2E and Supplementary Data**). While the corrected mutant cell fraction was calculated to be 85%, the gain of the mutant *CYSLTR2* allele was quantified to be present in 68% of the cells, giving clonal sizes of 17% and 68%, and a healthy cell fraction of 15% (**Figure 2F**). No copy number alterations involving chromosome 1p, 3p, 8q or 16q were detected (**Supplementary Table 2**).

In PUM-2B, we also observed two clones: in 83% of all cells the *CYSLTR2* p.L129Q mutation was identified, while 45% of the cells carried an extra copy of chromosome 13q. Accordingly, the two clones were present in 38% and 45% of the cells, and the remaining 17% represented healthy cells (**Figure 2F**).

The copy number profile (normal chromosomes 3p and 8) of this tumour is correlated to a relatively good prognosis, which is supported by the positive *BAP1* nuclear staining (**Supplementary Table 2**) (16). Indeed, after 8 years follow-up, no signs of metastatic spreading have been observed in this patient.

### Chromosome 13q alterations are uncommon in uveal melanoma

Since both *CYSLTR2* mutant tumours presented with chromosome 13q aberrations, we questioned whether such copy number alterations are common in *CYSLTR2* mutant and wild-type uveal melanomas. Therefore, we studied DNA sequencing and array genotyping data from 80 primary tumours from the TCGA (The Cancer Genome Atlas) cohort (13). 3/80 cases presented with a *CYSLTR2* p.L129Q mutation (V4-A9ED, VD-AA8O and YZ-A982), of which the clinical details are summarised in **Supplementary Table 2**. In two of these tumours many chromosomal alterations were identified, suggesting a complex polyploid genotype (**Figure S2**). However, a loss of the wild-type or gain of the mutant allele was not observed in these cases.

In the 77 *CYSLTR2* wild-type tumours, eight cases presented with a copy number alteration involving the *CYSLTR2* locus at chromosome 13q, making it an uncommon event in uveal melanoma (**Figure S3**).

### Mutant *CYSLTR2* is preferentially expressed at the RNA level

In the two *CYSLTR2* mutant uveal melanomas from our Leiden cohort, we also evaluated the gene expression of both wild-type and mutant allele by digital PCR. Although an allelic imbalance could be expected based on the genetic alterations identified, hardly any wild type allele expression was found, shifting the balance even more towards the mutant allele (**Figure 3A**). Despite a lower level of significance, preferential expression of the mutant allele was also observed in two of the three *CYSLTR2* mutant cases from the TCGA cohort (**Figure 3B**).

**Figure 3.**
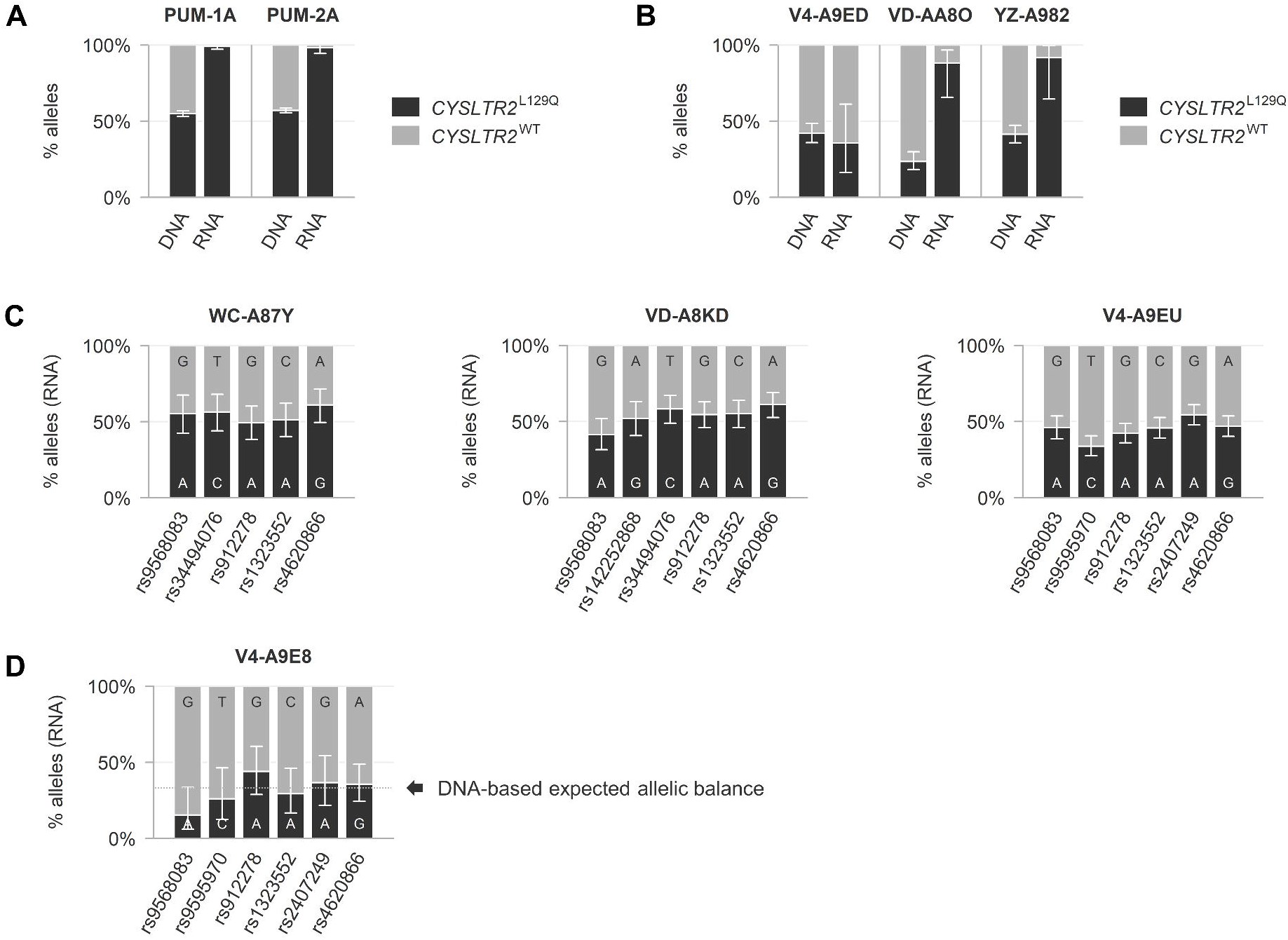
Allelic balance at the *CYSLTR2* locus in *CYSLTR2* mutant and wild-type uveal melanomas. (**A**) Comparison of DNA and RNA levels of mutant versus wild-type *CYSLTR2* alleles in two mutant uveal melanomas from our cohort, showing increased relative abundance of the mutant allele at RNA level in both tumours. (**B**) Three mutant uveal melanomas from the TCGA cohort, of which two show an allelic imbalance towards the mutant allele at RNA level. (**C**) Balanced biallelic expression of *CYSLTR2* in three copy number stable wild-type uveal melanomas from the TCGA cohort. (**D**) A wild-type uveal melanoma from the TCGA cohort with an allelic imbalance involving the *CYSLTR2* locus at chromosome 13q presents with proportional expression levels.

To gain insight into the regular transcriptional allelic balance of *CYSLTR2*, we studied RNA sequencing data from 77 *CYSLTR2* wild-type uveal melanomas from the TCGA cohort. The heterozygous presence of common SNPs illustrated a balanced biallelic expression under wild-type conditions (**Figure 3C**). In tumour V4-A9E8 a chromosome 13q copy number gain led to an allelic imbalance involving the *CYSLTR2* locus, which was proportionately expressed at the RNA level (**Figure 3D**).

It was however notable that the allelic balance of *CYSLTR2* could not be studied in all samples, as *CYSLTR2* expression varied largely between the melanomas (13).

### Wild-type *CYSLTR2* is expressed by melanoma cells in inflamed tumours

To investigate this variable *CYSLTR2* expression, we further analysed the 77 primary melanomas from the TCGA cohort and extended the analysis with the eight primary melanomas studied using single cell RNA sequencing as described by Durante et al. (13, 14). All these melanomas were wild-type for *CYSLTR2* and the majority carried mutations in *GNAQ, GNA11* or *PLCB4*.

Based on bulk RNA data from the TCGA cohort, the highest levels of wild-type *CYSLTR2* expression were observed in tumours with an inflammatory phenotype as shown by their immune gene expression signature (high expression of *CD14, CD163* [monocytes/macrophages] and *CD3D, CD8A* [T cells]) (**Figure 4A**). As *CYSLTR2* is known to be expressed by a variety of immune cells (12), we used the single cell RNA sequencing data from the Durante cohort to delineate the cellular origin of this wild-type *CYSLTR2* expression. In 5/8 cases *CYSLTR2* was expressed in at least 5% of the cells, with percentages of up to more than 30% (**Figure 4B**). Overall, the large majority of *CYSLTR2* expression originated from melanoma cells.

**Figure 4.**
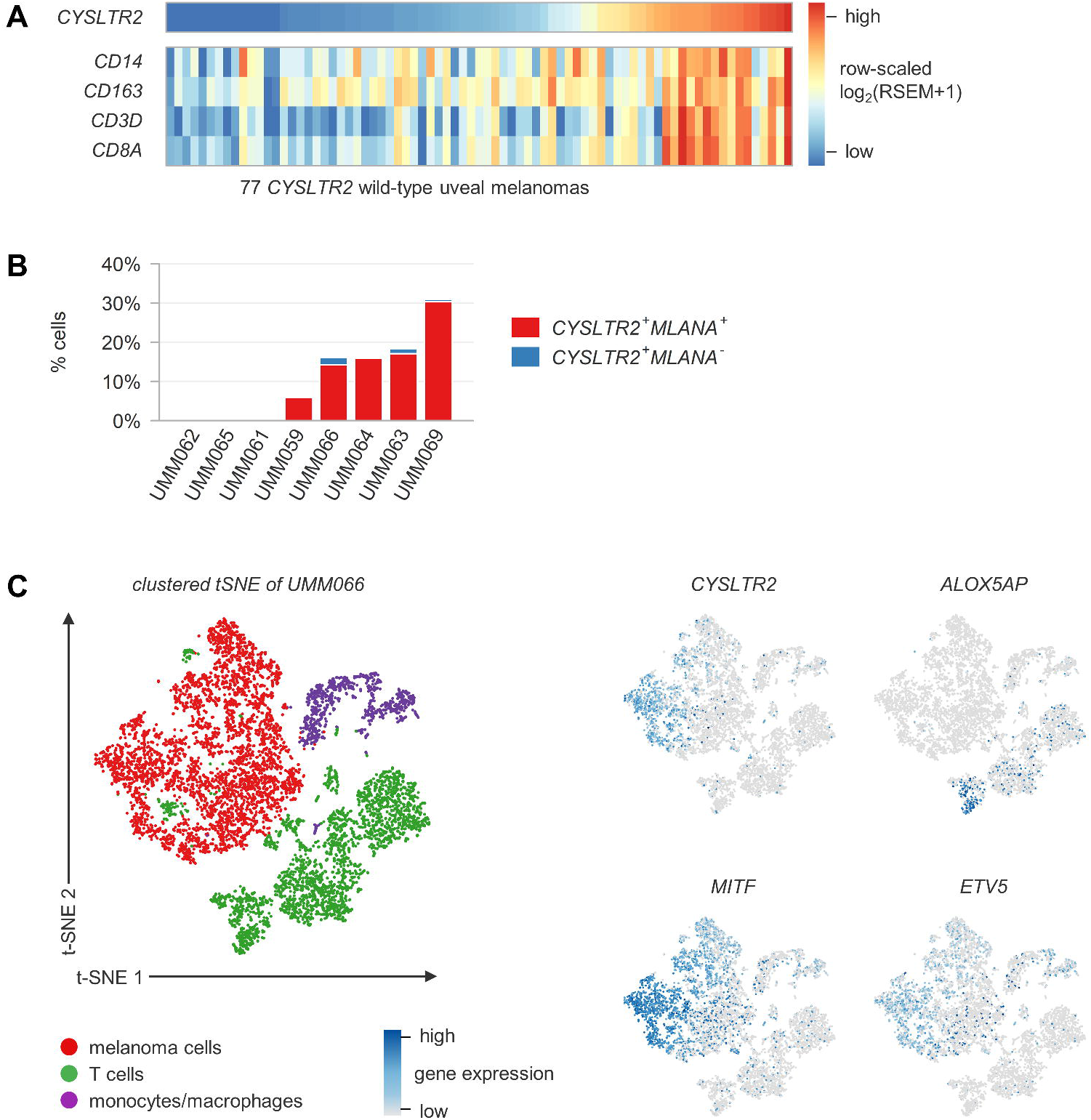
Origin of wildtype *CYSLTR2* expression in bulk uveal melanoma tissue and in single cell analysis. (**A**) Heatmap of row-scaled, normalised log_2_(RSEM+1) gene expression levels of *CYSLTR2, CD14* and *CD163* (monocytes/macrophages) and *CD3D* and *CD8A* (T cells) as determined by bulk RNA sequencing in 77 wild-type uveal melanomas from the TCGA cohort. The highest *CYSLTR2* expression levels are observed in the tumours with this immune gene signature expression. (**B**) Percentage *CYSLTR2* expressing cells per tumour annotated for cell type, as determined by single cell RNA sequencing in eight primary uveal melanomas from the Durante cohort. In 5/8 tumours *CYSLTR2* was expressed in >5% of the cells, which were predominantly melanoma cells. (**C**) tSNE plot of single cell RNA sequencing data derived from UMM066, with annotation of clusters for cell type and expression levels per cell of *CYSLTR2* (leukotriene receptor), *MITF* (pigmentation), *ETV5* (MAPK activation) and *ALOX5AP* (leukotriene synthesis). *CYSLTR2* was expressed in a subpopulation of melanoma cells that presented with high expression of genes involved in pigmentation and marker genes of MAPK activation. Enzymes catalysing the synthesis of leukotrienes (ligand of CysLT_2_R) are expressed by tumour-associated immune cells. Complete tSNE gene expression plots are presented in **Figure S4**.

As demonstrated in UMM066, *CYSLTR2* expression was found in melanoma cells that also presented with high expression of pigmentation genes *(MITF* and *TYR)* and marker genes of MAPK (Mitogen-Activated Protein Kinase) pathway activation (MPAS) (**Figure 4C and S4**) (21). As leukotrienes, the ligands of CysLT_2_R, cannot be measured at a single cell RNA-level, we used *ALOX5AP* and *ALOX5* as pseudomarkers (12). These genes, encoding enzymes catalysing the synthesis of leukotrienes, were specifically expressed by tumour-associated immune cells. Similar observations were made in the other tumours, suggesting that the immune infiltrate is equipped to produce the ligand for CysLT_2_R (**Figure S4**).

## Discussion

The majority of uveal melanomas are characterized by an activating Gα_q_ signalling mutation (4, 5). Whereas most tumours harbour a hotspot mutation in *GNAQ* or *GNA11*, the *CYSLTR2* p.L129Q mutation forms a rare alternative (9). In this study, we used digital PCR to analyse this mutation in non-malignant choroidal nevi and primary uveal melanomas. The potential role of wild-type *CYSLTR2* in *GNAQ, GNA11* or *PLCB4* mutant uveal melanomas was studied in publicly available bulk and single cell sequencing data.

Previously, mutually-exclusive mutations in *GNAQ* and *GNA11* (p.Q209L and p.Q209P) were found in 15/16 choroidal nevi (7). In the one remaining non-mutated nevus (no. 12B), we now identified *CYSLTR2* to be mutated (p.L129Q) (**Figure 1**). In the earlier study, another nevus from the same eye (no. 12A) showed a *GNA11* p.Q209L mutation (7). Taken together, these findings indicate that mutations in *GNAQ, GNA11* and *CYSLTR2* - all recurrently found in uveal melanomas - can also present as an independent somatic event in benign uveal nevi.

In cutaneous epidermal melanocytes, the *BRAF* p.V600E mutation - an established oncogenic driver of cutaneous melanoma - is considered sufficient to initiate nevus formation. This is supported by the frequent presence of this mutation in nevus cells (22, 23). The analysis of matched malignant and pre-malignant cutaneous lesions confirmed that a benign nevus may be a precursor of cutaneous melanoma (24). Likewise, it may now be hypothesised that Gα_q_ signalling mutations in *GNAQ, GNA11* and *CYSLTR2* represent a first step in uveal melanomagenesis.

We detected the *CYSLTR2* mutation in 2/120 (~2%) screened primary uveal melanomas. This low prevalence reaffirms the rarity of this variant (9). Both tumours in our study did not have a mutation in any of the hotspots of *GNAQ* or *GNA11*, confirming the mutual-exclusivity of these mutations.

Further analysis showed that the *CYSLTR2* p.L129Q was clonal in both of these uveal melanomas, supporting an acquisition of this mutation early during tumour development. However, in both melanomas secondary copy number alterations of the *CYSLTR2* locus (chromosome 13q) were observed as well: PUM-1 showed loss of the wild-type allele, while in PUM-2 the mutant allele was gained. These alterations were present in subclones of the melanomas, indicating that they arose later in the tumour evolution (**Figure 2**).

At the RNA level, the *CYSLTR2* allelic imbalance was skewed even further towards the mutant: in both melanomas hardly any expression of the wild-type allele was observed (**Figure 3A**). Although in one case (PUM-1) this may be attributed to the elimination of the wild-type allele during tumour progression, in the other case (PUM-2), a wild-type copy of *CYSLTR2* was still available. Similarly, in two of the three *CYSLTR2* mutant uveal melanomas from the TCGA cohort preferential expression of the mutant allele was observed, while in all cases the wild-type allele was still present (**Figure 3B**). This suggests that besides genetic mechanisms epigenetic mechanisms might also lead to an increased relative abundance of mutant *CYSLTR2*.

In *CYSLTR2* wild-type melanomas from the TCGA cohort, alterations involving chromosome 13q were uncommon and allelic balance was maintained in informative cases (**Figure 3C-D**). This supports that the *CYSLTR2* allelic imbalances at DNA and RNA levels are indeed associated with the p.L129Q mutation specifically. Whether these secondary alterations hold an additional metastatic risk, is - given the rarity of this mutation - still to be determined. However, the observation has been made that the mutation and secondary alterations involving *CYSLTR2* are found in all different molecular subtypes of uveal melanoma (**Supplementary Table 2**).

Again, our observations regarding *CYSLTR2* p.L129Q show similarities to *BRAF* p.V600E in cutaneous melanoma. Besides a primary role in driving the development of nevi and melanomas, secondary genetic or transcriptomic alterations are also recurrently present. Along with the progression from nevus to (metastatic) melanoma, absolute and relative copy-number increases and proportionally higher expression levels of the mutant allele are observed, enhancing oncogenic signalling (25). This possibly explains why therapies targeting mutant *BRAF* (a driver mutation already present in the pre-malignant cutaneous nevus) have been so successful in the treatment of metastatic melanoma (26). In analogy, mutant CysLT_2_R could be an attractive drug target for uveal melanoma. However, whereas effective antagonists for the other cysteinyl-leukotriene receptor (CysLT_1_R) are already in clinical use (i.e. montelukast), a novel compound functioning as an inverse agonist is needed to inhibit the constitutively activated mutant CysLT_2_R (9, 10, 27).

In bulk RNA from *CYSLTR2* wild-type uveal melanomas, we observed variable expression of *CYSLTR2*, with the highest levels in tumours with an immune gene expression signature (**Figure 4A**). These melanomas are characterised by a high infiltration of immune cells and are therefore seen as having an inflammatory phenotype (28, 29). Given that a variety of immune cells is known to express *CYSLTR2* (12), we investigated the cellular origin of this expression in these tumours. Using single-cell RNA data, we confirmed that wild-type *CYSLTR2* is mainly expressed by melanoma cells (**Figure 4B-C**). As subpopulations of tumour-associated immune cells expressed *ALOX5AP* and *ALOX5* (encoding the enzymes that catalyse the leukotriene synthesis), the ligand for wild-type CysLT_2_R may be produced by immune cells in the inflammatory microenvironment (**Figure 4C and S4**). We hypothesise that via this ligand, tumour-associated immune cells may interact with the uveal melanoma cells and further promote tumour growth.

Constitutive activation of the receptor (as seen with the p.L129Q mutation) leads to oncogenic signalling, cell proliferation and enforcement of a melanocyte-lineage-specific transcriptional program *in vitro* and *in vivo* (9, 10). Similarly, associations with a heavily pigmented phenotype have been reported in other melanocytic neoplasms carrying the p.L129Q mutation (30). It is interesting to speculate which effects leukotriene-driven activation of wild-type CysLT_2_R would have, as - based on our analysis of single cell data - wild-type *CYSLTR2* expressing cells presented with high expression of genes involved in pigmentation and MAPK pathway activation (**Figure 4C and S4**). Recently, it was shown *in vitro* that upon leukotriene activation wild-type CysLT_2_R can also signal through Gα_q_ and activate the MAPK pathway (10). Further research should evaluate whether such activation indeed has an extra oncogenic effect in these uveal melanoma cells which do not have a *CYSLTR2* mutation, but carry an activating Gα_q_ signalling mutation downstream in *GNAQ, GNA11* or *PLCB4*. As a consequence, wild-type CysLT_2_R may also represent a therapeutic target in uveal melanoma.

In contrast, mutant CysLT_2_R appeared unresponsive to leukotriene stimulation (10) and we observed that expression of the wild-type allele was downregulated in mutant tumours. This argues against a similar effect of the microenvironment in *CYSLTR2* mutant melanomas.

Importantly, the oncogenic alterations in uveal melanoma are not restricted to the *CYSLTR2* mutation. Therefore, we further studied the genetic heterogeneity of the two mutant melanomas from our cohort by digital PCR on multiple macro- and microdissected samples. In PUM-1 we found that the loss of wild-type *CYSLTR2* - together with the loss of chromosome 3p - was subclonally present and occurred after the *CYSLTR2* mutation, losses of chromosome 1p and 16q and gain of chromosome 8q. PUM-2 presented as a relatively stable tumour, but with a chromosome 13q alteration (leading to gain of mutant *CYSLTR2)* which also existed in a tumour subclone. In both melanomas the presence of the distinct clones was confirmed in spatially separated areas of the tumour, indicating that the clones coexist throughout different parts of the tumour (**Figure 2D and F**).

Hereby, we showed how the genetic heterogeneity and clonal evolution of a tumour can be construed from a deep quantitative analysis of bulk DNA, even in samples of lower DNA quality and quantity (i.e. FFPE-derived microdissections). The digital PCR experiments allowed us to analyse thousands of alleles per target, providing the statistical power to identify small allelic imbalances. Consequently, we identified differences in cancer cell populations which would not have been detectable using bulk sequencing-based techniques at conventional depths.

Whereas the genetic alterations in primary and - most recently - metastatic uveal melanoma have been largely catalogued, their relative order of occurrence in early tumoral development remains incompletely understood (31-33). Moreover, the existence of intra-tumoral heterogeneity in primary uveal melanoma is still a topic of debate (6, 14, 31-34). Our study exemplifies that these important questions may also be answered on bulk tissue samples by accurate quantitative analysis using digital PCR, presenting a useful addition to the single cell-based approaches (mass cytometry, single-cell sequencing) that are currently dominating the field.

## Conclusions

In conclusion, our findings strongly nominate the rare *CYSLTR2* p.L129Q mutation as an early oncogenic event in *GNAQ* and *GNA11* wild-type uveal nevi and melanomas. In mutant melanomas, the relative gene dosage of the *CYSLTR2* mutant allele may be further increased during progression of the primary tumour. This makes these tumours promising candidates for mutant CysLT_2_R-targeted therapy.

Moreover, we identified the expression of wild-type *CYSLTR2* in inflamed tumours carrying a mutation in *GNAQ, GNA11* or *PLCB4*. As this expression originated from the melanoma cells specifically, wild-type CysLT_2_R also needs to be considered as a potential therapeutic target.

## Data Availability

The data sets analysed during the current study are available via the Broad GDAC Firehose, NCI Genomic Data Commons data portal (TCGA Uveal Melanoma data set) and Gene Expression Omnibus repository (Durante et al. single cell RNA-data set, GSE139829).

https://gdac.broadinstitute.org)

https://portal.gdc.cancer.gov

https://www.ncbi.nlm.nih.gov/geo/query/acc.cgi?acc=GSE139829

**List of abbreviations**
FFPE: Formalin-Fixed Paraffin-Embedded
LUMC: Leiden University Medical Center
MAPK: Mitogen-Activated Protein Kinase
PCR: Polymerase Chain Reaction
PUM: Primary Uveal Melanoma
SNP: Single Nucleotide Polymorphism
TCGA: The Cancer Genome Atlas

## Declarations

### Ethics approval and consent to participate

16 choroidal nevi were dissected from 13 formalin-fixed post-mortem human donor eyes, with consent from Lions NSW Eye Bank and approval from the University of Sydney and University of New South Wales Human Research Ethics Committee (7). 120 fresh-frozen tumour samples, obtained from primary uveal melanomas treated by enucleation, were available from the Biobank of the Department of Ophthalmology, LUMC. All patient samples were collected with informed consent and this study was approved by the LUMC Biobank Committee and Medisch Ethische Toetsingscommissie under no. B14.003/DH/sh and B20.026.

### Consent for publication

All patients samples were collected with informed consent. All authors agree to the content of the paper and their being listed as a co-author.

### Availability of data and materials

The data sets analysed during the current study are available via the Broad GDAC Firehose (https://gdac.broadinstitute.org), NCI Genomic Data Commons data portal (GDC; https://portal.gdc.cancer.gov) and Gene Expression Omnibus repository (https://www.ncbi.nlm.nih.gov/geo/query/acc.cgi?acc=GSE139829) (13, 14).

### Authors’ contributions

RJN and PAvV conceptualised this study and wrote the manuscript. MV, GPML, RMV, MCM and MJJ collected the patient materials. RJN, NVM and MV performed the PCR experiments, RJN performed the bioinformatic analyses, and RMV and MCM performed the histological examination. MV, RMV, MCM and MJJ revised the manuscript. PAvV supervised this study. All authors read and approved the final manuscript.

## Acknowledgements

We thank Nelleke A. Gruis (Department of Dermatology, LUMC) and Astrid G.S van Halteren (Department of Pediatrics, LUMC) for helpful discussions and reviewing the manuscript.

## Supplementary information

**Supplementary Table 1**

Context sequences, PCR annealing temperatures and supplier information for all assays used.

**Supplementary Table 2**

Clinical and molecular characteristics of *CYSLTR2* mutant uveal melanomas (PUM-1, PUM-2, V4-A9ED, VD-AA8O and YZ-A982) analysed in this study. Part of the information has been obtained from uncontrolled, publicly available data from the original TCGA study on uveal melanoma (13).

**Supplementary Data 1**

Methodical overview of all digital PCR experimental setups.

**Supplementary Figure 1**

*CYSLTR2* p.L129Q mutation detected by Sanger sequencing of choroidal nevus 12B compared to the reference sequence from a healthy control.

**Supplementary Figure 2**

SNP-array copy number profile and B-allele fractions of the *CYSLTR2* mutant melanomas from the TCGA cohort.

**Supplementary Figure 3**

Distribution of arm-level copy number alterations in 77 *CYSLTR2* wild-type uveal melanomas from the TCGA cohort.

**Supplementary Figure 4**

Origin of *CYSLTR2* expression and characterization of the immune microenvironment in uveal melanoma based on the single cell RNA expression analysis of the five tumours with >5% *CYSLTR2^+^* cells.

